# Hepatitis B virus infection in the Lao PDR: A systematic review

**DOI:** 10.1101/2022.01.23.21265872

**Authors:** Lisa Hefele, Phonethipsavanh Nouanthong, Judith M. Hübschen, Claude P Muller, Antony P Black

**Affiliations:** Department of Infection and Immunity, Luxembourg Institute of Health, Esch-sur-Alzette, Grand-Duchy of Luxembourg; Lao-Lux Laboratory, Institut Pasteur du Laos, Ban Kao-Gnot, Sisattanak district, Vientiane, Lao PDR

**Keywords:** Hepatitis B, Public Health, Epidemiology

## Abstract

**Introduction:** Even though hepatitis B is endemic in the Lao PDR, the understanding of the epidemiology of hepatitis B infection is incomplete. This article reviews the available literature about hepatitis B seroprevalence, risk factors and genotypes in the Lao population in order to provide an up- to date summary of the HBV epidemiology in the Lao PDR, identify knowledge gaps and provide public health recommendations.

**Methods:** Using PubMed/Medline and ScienceDirect, all studies reporting the prevalence of hepatitis B markers or genotype distribution in the Lao PDR published were systematically reviewed.

**Results:** The 21 studies included focused on the general population, blood donors, women, children, health care workers and garment factory workers. The studies varied extensively in sample size, target population, methods, study location and time periods. The prevalence of the hepatitis B surface antigen (HBsAg) in blood donors was reported to be 8.7%-9.6% in 2003-2006. In the years 2011-2012, the reported HBsAg prevalence among women (including pregnant women) ranged from 0%-9.5% and among children aged 5-9 years from 1.7%-8.7%, depending on study location and age. The majority of strains characterized in Lao PDR belonged to genotypes B and C.

**Conclusion:** Studies displayed considerable heterogeneity in populations, design and laboratory methods. A high HBsAg prevalence was reported in adults including pregnant women. Low infant vaccination coverage and compromised vaccine immunogenicity were found. Only two studies focused on HBV in risk populations, emphasizing the need for further studies to characterize hepatitis B epidemiology in potentially vulnerable groups. Hepatitis B infection continues to represent a substantial public health threat in the Lao PDR and needs to be monitored to inform health authorities and to counteract over-burdening of the health care system. In order to end mother to child transmission, vaccination coverage with the hepatitis B birth dose should be increased.

## Introduction

Hepatitis B and C, responsible for 96% of all hepatitis mortality, cause more deaths than either HIV/AIDS, tuberculosis or malaria and yet seem to receive much less attention. The WHO African and Western Pacific Regions are the most affected by the Hepatitis B virus (HBV). Although most infections with HBV are asymptomatic, in some patients, acute infection can cause severe liver inflammation. Chronic infection with HBV can lead to liver cirrhosis and hepatocellular carcinoma (1,2). The Coalition for Global hepatitis elimination estimated that globally 40% of liver cancer deaths could be attributed to HBV and the global number of HBV-related deaths was estimated at more than half a million in 2019 (3).

HBV is grouped into at least 9 recognized genotypes (A to I), and one presumable genotype “J”, each with a distinct geographic distribution. Evidence suggests that the HBV genotypes are related to HBV transmission modes and clinical outcomes (4–7). In Asia, genotypes B and C dominate largely (8,9) and are associated with vertical transmission (4).

Even though HBV is recognized as a major public health threat in the Lao PDR, it is challenging to provide quantitative epidemiological data about the HBV prevalence in the country. This is due to several reasons, including an inadequate surveillance system (10), the geographic and ethnic heterogeneity of the country and limited HBV testing. Understanding the HBV epidemiology in Lao PDR is important for improving vaccination, testing and treatment strategies.

Routine vaccination for children is free of charge in Lao PDR. Since 2001, HBV containing vaccines were gradually introduced in the routine national immunization schedule at 6, 10 and 14 weeks of age. Currently, HBV vaccination is administered as part of the pentavalent routine infant vaccine in combination with diphtheria, tetanus, pertussis and *Haemophilus influenzae* type b (DTPw-HepB-Hib). Vaccination coverage rates vary regionally, but the nationwide coverage of 3 doses of the DTPw-HepB-Hib reached 84% in 2018 (11). The monovalent HBV birth dose was introduced in Lao PDR in 2003 (12,13), but coverage rates were only 55% in 2018 (11). The low coverage rates for the birth dose are particularly concerning since mother-to-child transmission (MTCT) is one of the main routes of transmission in endemic countries (14). To date, only pregnant women are routinely screened for HBV during antenatal care visits, but immunoglobulin and anti-viral treatment are not readily available in the Lao PDR. The high prevalence of HBV infections in the adult Lao population contributes to the high levels of liver cancer (22.4 per 100,000; 5^th^ highest worldwide (15)) and will likely continue to perpetuate there.

Despite several studies in Lao PDR, our insights in the epidemiology of Hepatitis B in the country are still fragmented. This systematic review, the first in Lao PDR, will provide an up-to-date summary of the HBV situation in the country. We aim to address the following questions: which data are available? Which conclusions can be drawn for national prevention and intervention strategies? What are the specific modalities of HBV transmission or susceptibility in particular risk groups? What are the prevalent HBV genotypes in the Lao PDR? What are current knowledge gaps?

## Methods

### Literature review and study selection

A systematic review of peer-reviewed literature on HBV in the Lao PDR was conducted following the PRISMA statement guidelines (16). PubMed and ScienceDirect (Elsevier) were searched by using the following key words: “Lao PDR” (or “Laos” or “Lao People’s Democratic Republic”) in combination with “hepatitis B” (or “HBV”), resulting in the full search term “((“hepatitis B”) OR (“HBV”) OR (“jaundice”)) AND ((“Lao PDR”) OR (“Lao People’s Democratic Republic”) OR (“Laos”))”. The last day of search was 16.04.2021.

All articles were managed with the bibliographic management tool EndNote. Duplicate entries were removed. All peer-reviewed studies reporting prevalence of HBV infection or HBV genotype distribution in the Lao PDR and published between 01.01.1990 and 16.04.2021 were included. Reports were screened and all reports which did not meet the eligibility criteria or were reviews, book chapters, case reports, or conference abstracts were removed (Fig 1).

**Fig 1.**
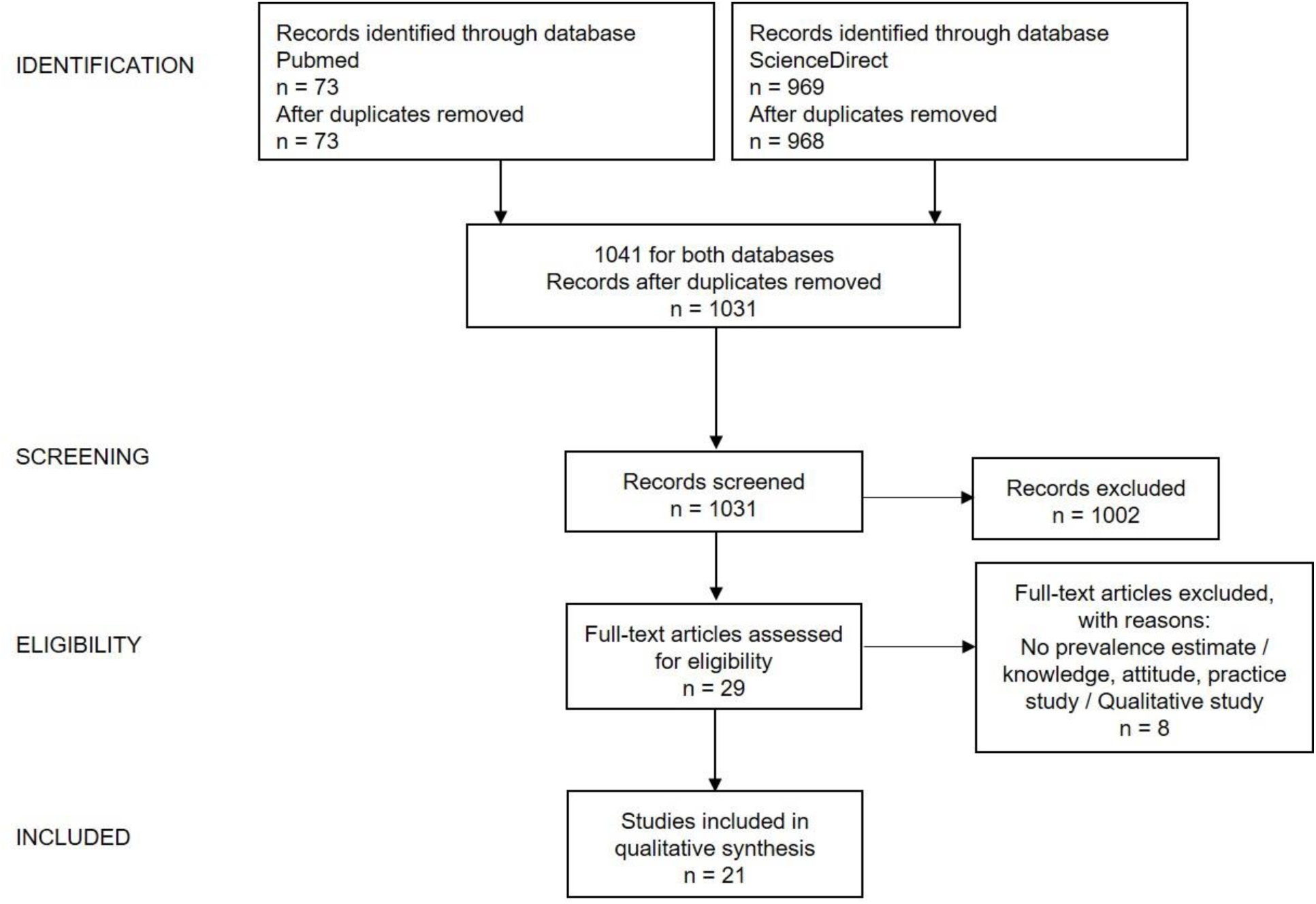
Flow diagram of the systematic review regarding Hepatitis B genotypes and Hepatitis B prevalence in the Lao PDR, 1990-2021.

### Data analysis

After selection of eligible studies, the following study data were extracted and entered into a table: study identification data (first author, publication year and journal of publication), study period, location, population, design, age of participants and estimates of prevalence of HBV markers or HBV genotype distribution. Prevalence of the hepatitis B surface antigen (HBsAg) was defined as the main primary outcome in addition to the prevalence of other HBV markers indicating past infection (anti-Hepatitis B core antigen antibodies; anti-HBc) or past infection/vaccination (anti-Hepatitis B surface antigen antibodies; anti-HBs). HBV genotype distribution was included as secondary outcome. Due to the scarcity of HBV prevalence studies in the Lao PDR and the heterogeneity of available studies, limitations and possible study biases were discussed for each study individually.

## Results

### General scope

Of the 1031 identified references, 1002 were excluded after screening the title or/and abstract. Of the remaining 29 articles, 21 were deemed eligible for inclusion in the systematic review (Table 1).

**Table 1.** Overview over all articles included in the review.

| Year | Authors | Title |
| --- | --- | --- |
| 1998 | Bounlu et al. | Acute Jaundice in Vientiane, Lao People's Democratic Republic |
| 2007 | Jutavijittum et al. | Seroprevalence of Hepatitis B and C virus infections among Lao blood donors |
| 2008 | Olinger et al. | Possible new hepatitis B virus genotype, Southeast Asia |
| 2010 | Syhavong et al. | The infective causes of hepatitis and jaundice amongst hospitalised patients in Vientiane, Laos |
| 2012 | Andernach et al. | A High Variability of Mixed Infections and Recent Recombinations of Hepatitis B Virus in Laos |
| 2013 | Jutavijittum et al. | Occult hepatitis B infections among blood donors in Lao PDR |
| 2014 | Black et al. | Hepatitis B virus in the Lao People's Democratic Republic: a cross sectional serosurvey in different cohorts |
| 2014 | Xeutavongsa et al. | Chronic Hepatitis B Prevalence among Children and Mothers: Results from a Nationwide, Population-Based Survey in Lao People's Democratic Republic |
| 2015 | Black et al. | Serosurveillance of Vaccine Preventable Diseases and Hepatitis C in Healthcare Workers from Lao PDR |
| 2015 | Komada et al. | Seroprevalence of chronic hepatitis B, as determined from dried blood spots, among children and their mothers in central Lao People's Democratic Republic: a multistage, stratified cluster sampling survey |
| 2016 | Jutavijittum et al. | High rate of Hepatitis B virus mother-to-child transmission in Lao People's Democratic Republic |
| 2016 | Xaydalasouk et al. | Assessment of mother-to-child HBV transmission at the prenatal consultation in Vientiane, Laos |
| 2016 | Evdokimov et al. | Low and disparate seroprotection after pentavalent childhood vaccination in the Lao People's Democratic Republic: a cross-sectional study |
| 2017 | Choisy et al. | Prevalence of Hepatitis B Virus Infection among Pregnant Women Attending Antenatal Clinics in Vientiane, Laos, 2008–2014 |
| 2018 | Xaydalasouk et al. | Seroprevalence and risk factors of hepatitis B and C virus infections in female workers of Lao garment factories |
| 2019 | Hefele et al. | Seroprotection at different levels of the health care system after routine vaccination with DTPw-HepB-Hib in Lao PDR |
| 2019 | Norizuki et al. | Serologic testing of randomly selected children after hepatitis B vaccination: a cross-sectional population-based study in Lao People's Democratic Republic. |
| 2019 | Latthaphasvan et al. | Perinatal hepatitis B virus transmission in Lao PDR: A prospective cohort study |
| 2020 | Hefele et al. | Lasting benefit of infant hepatitis B vaccination in adolescents in the Lao People's Democratic Republic |
| 2020 | Mangkara et al. | Hepatitis B virus in Lao dentists: A cross-sectional serological study |
| 2021 | Xaydalasouk et al. | Age-stratified seroprevalence of vaccine-preventable infectious disease in Saravan, Southern Lao People's Democratic Republic. |

From the three articles which investigated the genotype distribution of HBV in the Lao PDR (17–19), two studies also reported HBsAg prevalence (18,19). The remaining 18 articles focused solely on the prevalence of HBV markers. Four studies were conducted in first-time blood donors (17–20), two studies investigated the aetiology of jaundice and hepatitis in Vientiane (21,22), 12 studies were conducted in the general population (including pregnant women) (12,23,32,33,24–31) and three studies focused on risk groups (34–36).

In terms of study design, almost all studies were cross-sectional (n=17), one study was a retrospective analysis of medical records, one study was an aetiological study, one study was a case-control study and one study was a prospective study.

### Patients with acute jaundice

An early report investigated the aetiology of acute jaundice in three hospitals in Vientiane in 1995-1996 (22) (Table 2). In this study, HBV was identified as one of the major causes of acute viral hepatitis (10% IgM anti-HBc), the most common being Hepatitis A (14%). HBV was also identified as an important cause of hepatitis or jaundice in hospitalized patients in Vientiane from 2001 to 2004 (21).

**Table 2.**
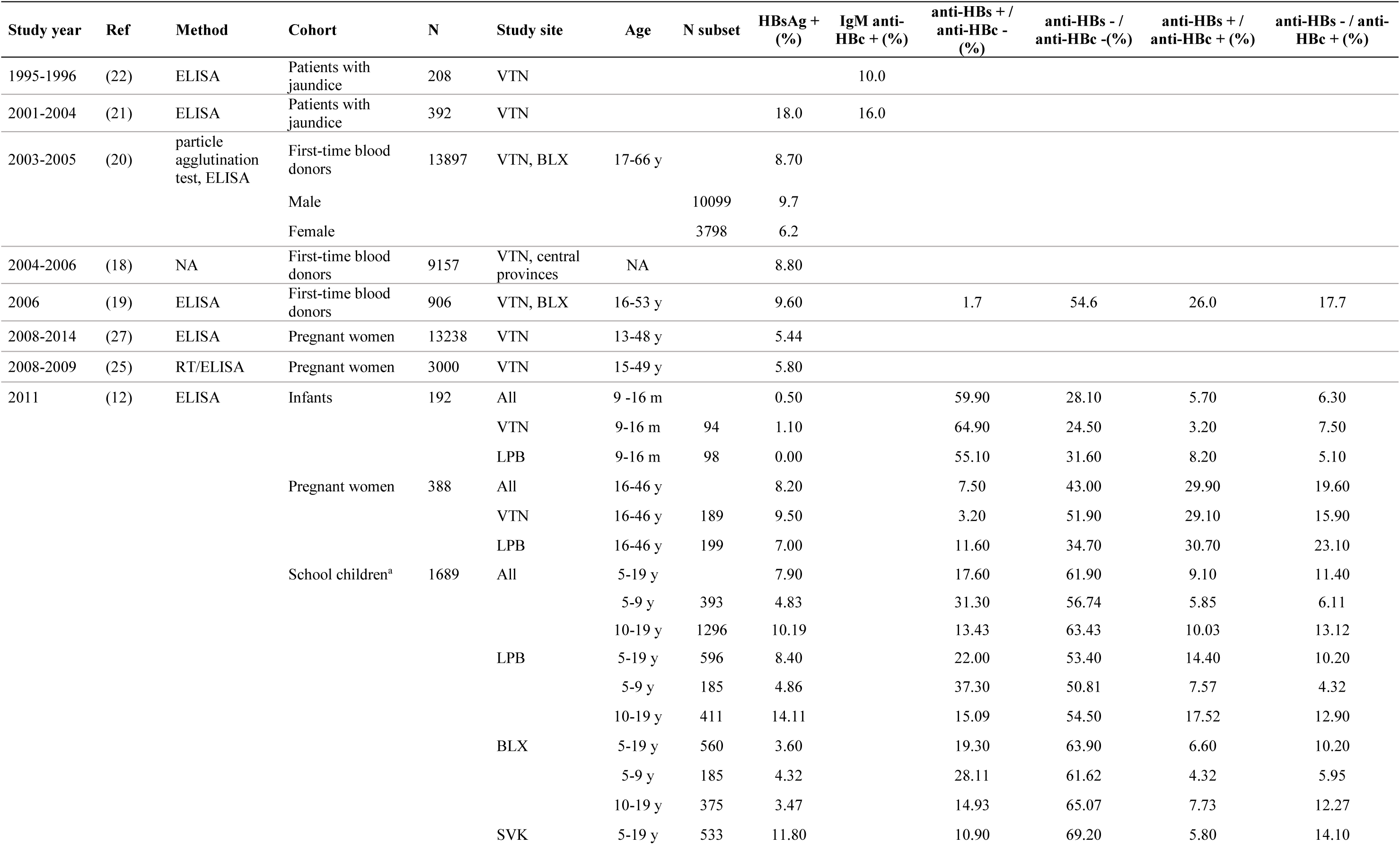
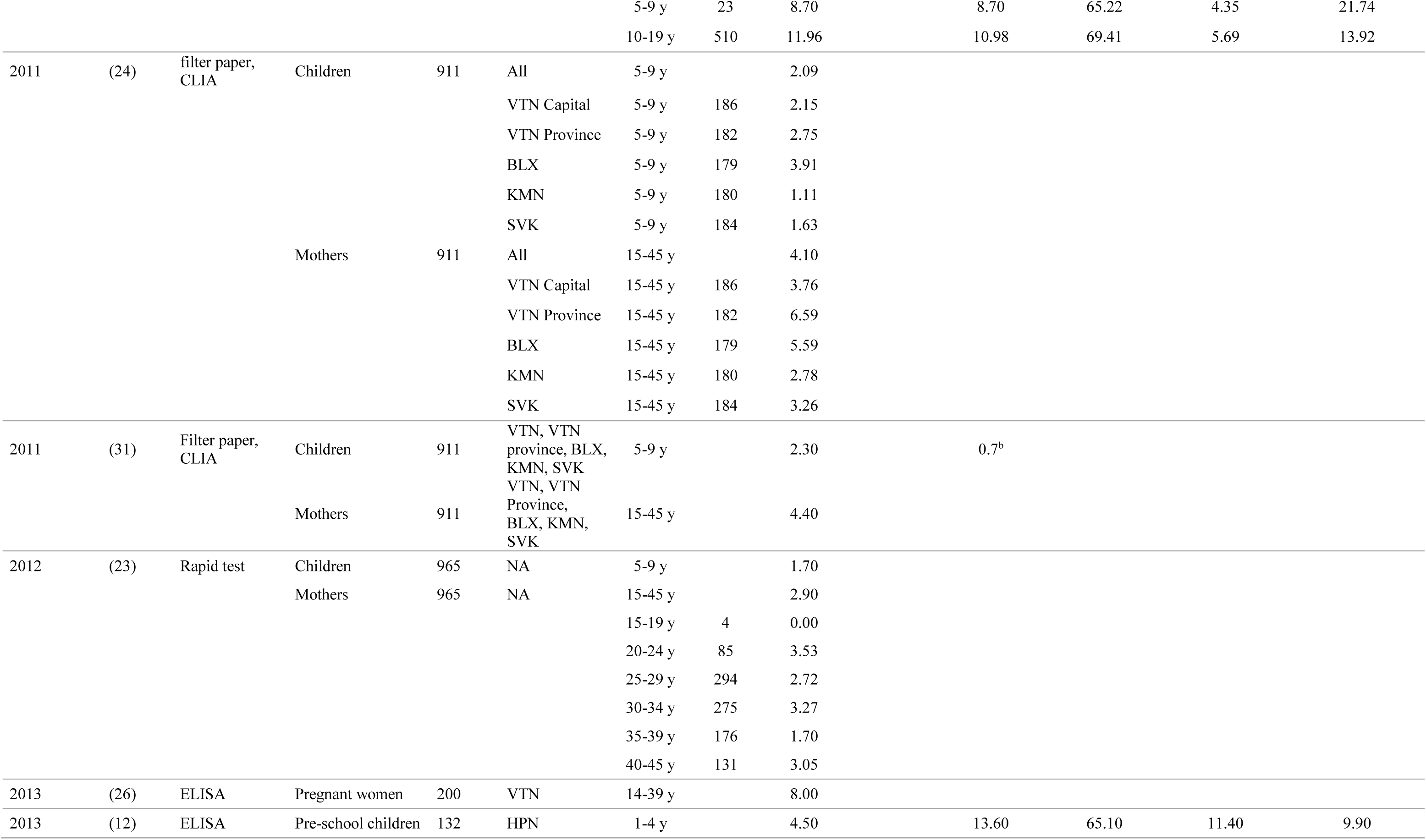
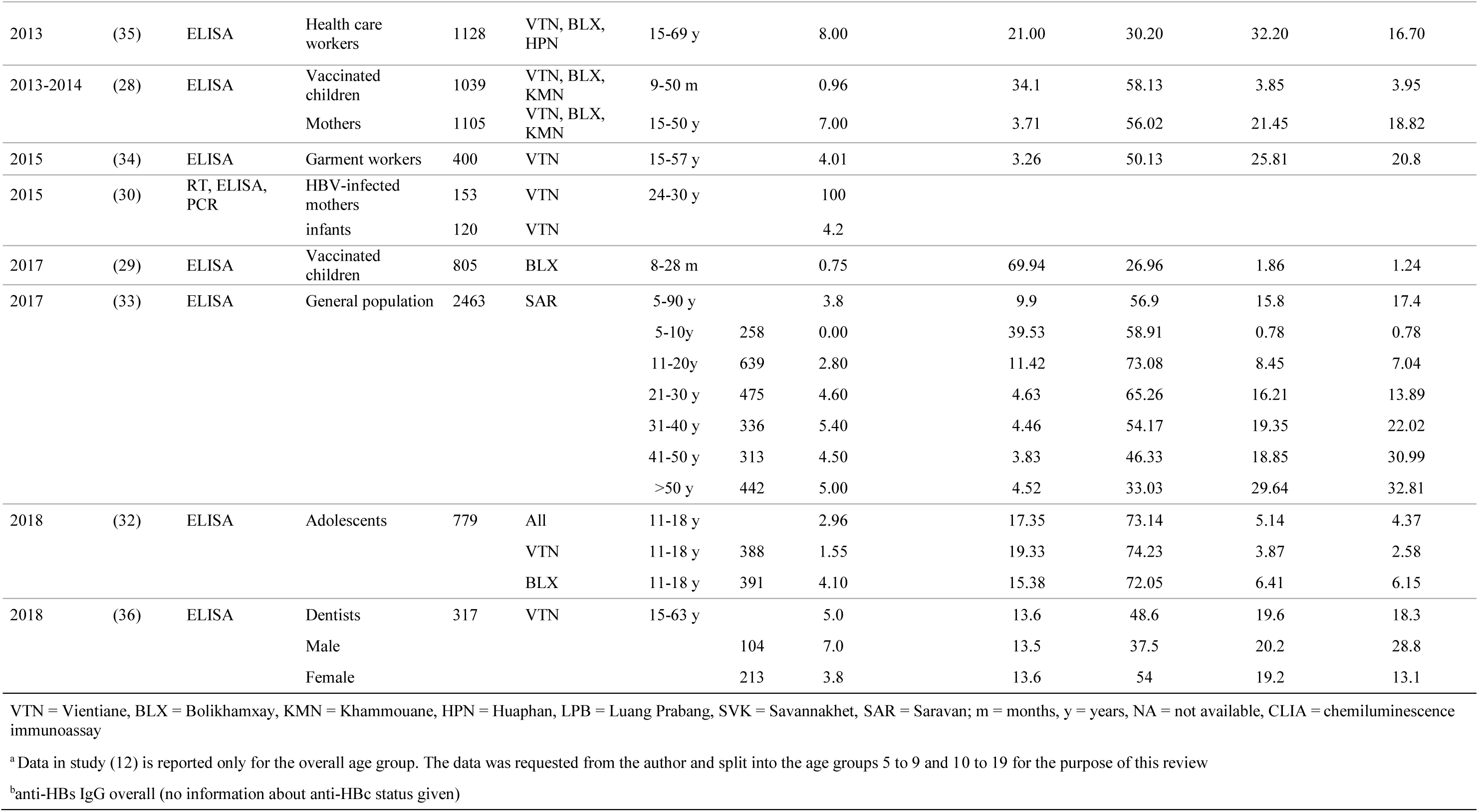
Data regarding HBV prevalence in the Lao PDR.

### HBV seroprevalence

The 18 seroprevalence studies varied in sample size, target population, methods, location and time periods (Table 2). All studies but one provided HBsAg estimates and 9 studies also investigated the prevalence of anti-HBs and anti-HBc antibodies.

#### General population

Most seroprevalence studies assessed the HBV seroprevalence in women and children; however, one cross-sectional study in Saravan province also reported HBV prevalence in men.

Reported HBsAg prevalence estimates for women ranged from 0 to 9.5% in the general population (Table 2).

The earliest estimate of 6.2% chronic HBV infections in women was from the above study focusing on blood donors between 2003 and 2005 (20). Similarly, a hospital-based study in Vientiane from 2008 to 2009 found 5.8% of pregnant women aged 15-49 years to be HBsAg positive (25) (Table 2, Fig 2).

**Fig 2.**
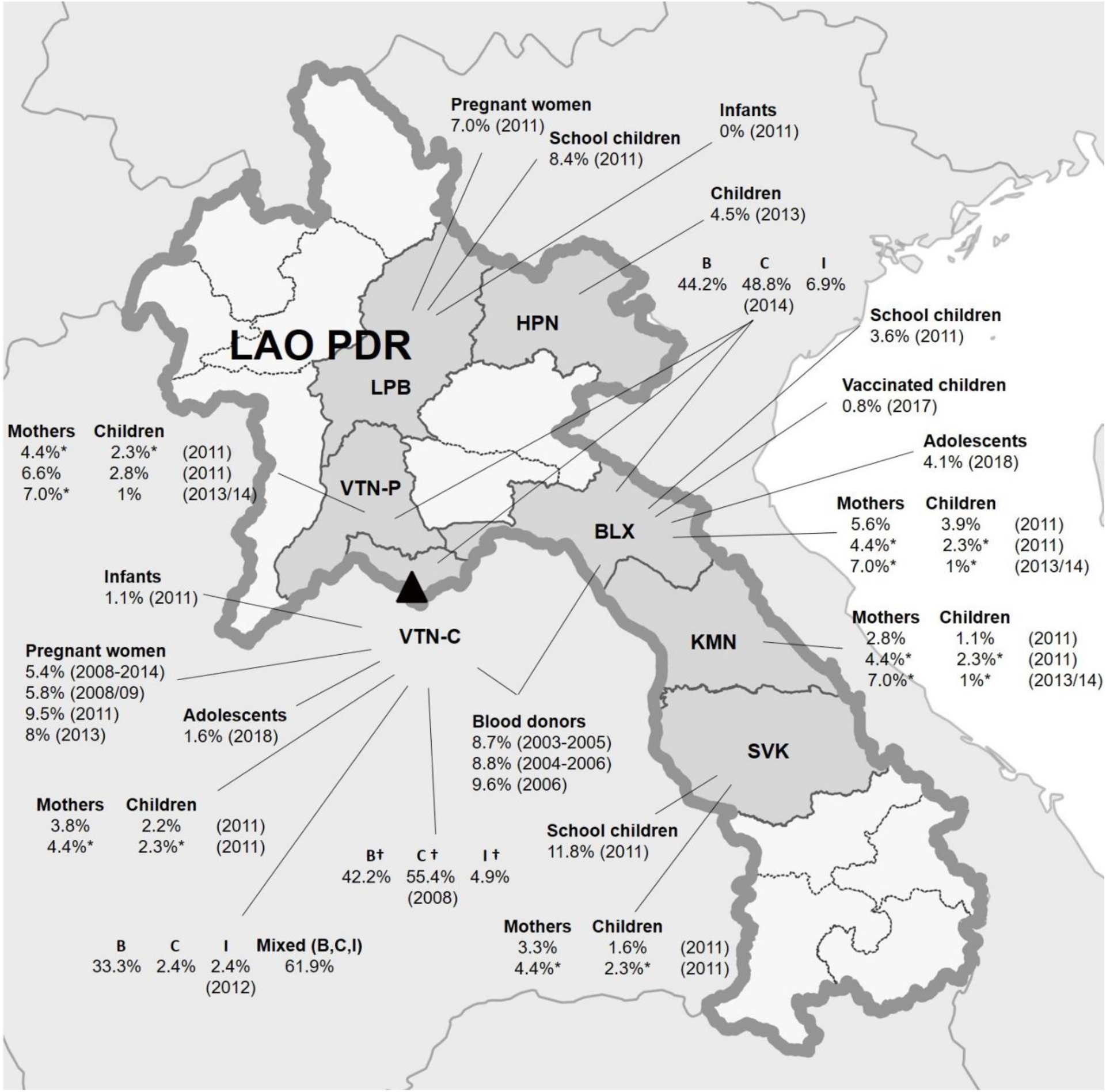
Geographical distribution of HBsAg prevalence in the general population and HBV genotypes in the Lao PDR. HBsAg prevalences in sub-populations are depicted per province (if information was given in the article), with the study year in brackets. * = HBsAg estimate was given as an overall estimate of several study locations. † = study location in article stated as Vientiane Capital and Central provinces. The location of Vientiane Capital is indicated by the black triangle. LPB = Luang Prabang, HPN = Huaphan, VTN-P = Vientiane Province, VTN-C = Vientiane Capital, BLX = Bolikhamxay, KMN = Khammouane, SVK = Savannakhet, SAR = Saravan.

Between 2011 and 2012, four other studies characterized the HBV prevalence in women and children (12,23,24,31).

In 2011, a stratified, multistage, cluster sampling survey was conducted in central Lao PDR including 911 mother-child pairs (24). The overall HBsAg seroprevalence was 4.1% in the mothers (15-45 years) (Table 2, Figure 2). Estimated HBsAg prevalence rates differed between study sites in Vientiane municipality (3.8%), Vientiane province (6.6%), Bolikhamxay (5.6%), Khammuane (2.8%) and Savannakhet (3.3%).

In a similar study in mothers (15-45 years) and their children in the same central provinces in the same year (2011), (31) found an HBsAg prevalence of 4.4% in the mothers.

A non-randomized, cross-sectional seroprevalence study including multiple cohorts (12) reported 9.5% and 7% HBsAg rates in pregnant women (16-46 years) in Vientiane and Luang Prabang, respectively, and nearly half of all pregnant women were positive for anti-HBc (49.5%).

In 2012, in another multistage, cluster sampling mother-child survey, HBsAg prevalence was only 2.9% in mothers (15-45 years) (23), but there was no indication of the specific study sites within Lao PDR.

Tree of the mother-child studies (23,24,31) used cluster sampling to estimate HBsAg prevalence rates in the Lao PDR, but they differed in their testing methodology. In two studies (24,31), samples were collected onto filter paper via finger prick and analysed by chemiluminescence immunoassay while in another study (23), a rapid test was used to determine HBsAg status. The authors of one study (24) remarked in their Discussion on issues regarding the utilization of dried blood spots in HBV serosurveys since there are not many reports investigating the diagnostic reliability in the context of field work.

A non-randomized cross-sectional seroprevalence study conducted in 2013/14 in Khammouane, Vientiane and Boulhikhamxay provinces (28) also recruited mother-child pairs, but focused on children with a complete course of the DTPw-HepB-Hib vaccine. Overall HBsAg prevalence in mothers was 7%, higher than the overall HBsAg prevalence reported in mothers by some of the above studies (23,24,31), indicating regional variation or possibly different sensitivity and specificity of the techniques used for assessing HBsAg positivity.

A study conducted in 2013 in Vientiane capital reported the HBsAg seroprevalence to be 8% in pregnant women (14-39 years) (26). Another retrospective study during 2008-2014 described HBsAg prevalence among pregnant women visiting the Mahosot hospital in Vientiane as 5.4% (27). A slight yet steady decline of the prevalence was reported over the period of the study. These rates in pregnant women were somewhat higher than the prevalence of HBsAg reported in women in two of the mother-child studies (23,24).

The cross-sectional study conducted in Saravan reported 33.2% of the total tested population to have anti-HBc antibodies indicating previous exposure to HBV. The proportion of exposed individuals increased with age and the proportion of exposure in males was significantly higher than in females (37.4% and 30.0%; p<0.0001). Overall, 5% of the general population were chronically infected(33).

#### Children and adolescents

MTCT is one of the main routes of transmission in endemic countries (14). The hospital-based study in Vientiane in 2008/09 (25) which found 5.8% pregnant women aged 15-49 years to be HBsAg positive reported that as many as 21% of infants born to these HBsAg positive mothers were also HBsAg positive (Table 2). Unfortunately, the vaccination history of the children was not recorded in this study and it is unknown if they had received the HBV birth dose, which was introduced in 2003.

MTCT was investigated in a prospective-cohort study on HBsAg positive pregnant women and their new-borns from 2015-2017 in Vientiane Capital (30). HBV vaccination was provided for all new-borns according to the immunization schedule. The authors reported a relatively low rate of MTCT considering that Hepatitis B immunoglobulin and anti-viral treatment are not readily available in the Lao PDR: 4% of children born to chronically infected mothers were positive for HBsAg. 15 of the 120 infants showed unsatisfactory protective antibody levels after vaccination when tested at 6 months of age (anti-HBs <10 IU/L), indicating the need for improving vaccine management.

The three multistage cluster sampling studies (23,24,31) found an overall HBsAg seroprevalence between 1.7% and 2.3% among 5 to 9 year old children (Table 1). Again, the HBsAg prevalence reported by another study (12) was more than twice as high (4.8%) in children of the same age range.

In a study from 2011, (31), participants were tested for both HBsAg and anti-HBs. The study reported that only 0.7% of all the included children had a positive anti-HBs titer (with or without documented vaccination) approximately 3 to 9 years after being presumably vaccinated. None of the children with a documented vaccination history showed protective anti-HBs titers (here defined as 10 UI/L).

A separate study (12) reported substantial heterogeneity in terms of vaccination coverage and health care access in the Lao PDR in 2011. Four sub-populations from different age groups were investigated regarding the prevalence of HBV markers. While infants showed a low prevalence of HBsAg (0.5%) and a relatively high rate of the serological vaccination profile (59.9%), school children born before and after the introduction of the vaccine in 2001 had a much higher HBsAg prevalence (7.9%). In 2011, school children (age 5 to 9 years) were included in this study (12) and one mother-child study (24) from the same location. While the prevalence of HBsAg in these school children reported from Bolikhamxay was similar in both studies (3.6% and 3.9%), the mother-child study (24) reported only 1.63% of the children to be HBsAg positive in Savannakhet, in contrast to the school children study (12), which found 8.7% of the children to be infected. Reasons for the discrepancy in HBsAg prevalences are not clear, but the studies differed greatly in design and methodology. In the mother-child study (24), participants were selected randomly, samples were collected onto filter paper via finger prick and analysed by chemiluminescence immunoassay while in the school children study (12), recruitment took place in the framework of a national measles and rubella vaccination campaign, samples were taken by venipuncture and analysed by enzyme-linked immunosorbent assay.

Although HBV vaccination was introduced in the national immunization schedule in 2001, only 13.6% of pre-school children (1-4 years) from a rural area in Huaphan province in Northern Lao PDR showed the serological vaccination profile in 2013 and 4.5% were positive for HBsAg. This indicated that vaccine coverage was very low in this setting even 10 years after the introduction of the vaccine (12) (Table 2, Fig 2).

In a study in 2013/14 less than 1% of children aged 9-50 months with documented three doses of DTPw-HepB-Hib were chronically infected, despite a low proportion of children showing a serological vaccination profile (34%) (28). These results show that in addition to low vaccine coverage, vaccine immunogenicity is a problem in the Lao PDR. A follow-up study in 2017 reported a considerable improvement in vaccine immunogenicity and a decrease in HBV infection rates: HBsAg prevalence declined from 1.8% to 0.8%, when only children in the same age range were compared between 2013/14 and 2017 (29). Results indicated significantly lower protection rates in remote areas.

In 2018, a cross-sectional seroprevalence study investigated the impact of the HBV vaccination in central Lao PDR in randomized adolescent school children from 11 to 18 years, born just before or after the infant vaccine introduction (32). In this study, the proportion of students with a serological vaccination profile increased after the introduction of HBV vaccine and a sizable number were still protected 11 years after vaccination. In addition, the prevalence of past infection decreased.

#### Risk groups

A study conducted in 2013 revealed a very high proportion of health care workers to be susceptible to HBV (35). Most participants included in the study were female (79.8%). 8% were HBsAg carriers (11.5% in males and 7.1% in females) and about half (48.8%) had anti-HBc antibodies indicating previous exposure (Table 2). The HBsAg prevalence of 7.1% in female health care workers was higher than reported for women in some studies (23,24,31), but is comparable to others (12,26,28). Dental workers in a study from 2018 also showed a high proportion of previous exposure (37.8% anti-HBc+) and 5% were chronically infected(36).

A study in 2014 focused on female garment factory workers aged 15 to 57 years as a vulnerable population (34). Although the HBsAg prevalence of 4% was lower than expected (Table 2), the data suggested a significant association with sexual risk behaviour and very low levels of knowledge and awareness regarding HBV. More than 40% of the women showed evidence of past infection with HBV.

### Risk factors or predictors associated with HBV infection

Although some studies found no risk factors associated with HBV infection status (37), others have identified significant associations. HBsAg prevalence among blood donors was higher in males and increased significantly with age (19,20). In Saravan province, previous exposure to HBV was significantly higher in males than females and increased with age (33). Chronic infection in the mothers was a risk factor for chronic infection in children aged 5-9 years in two studies (23,24). Being born outside a health care facility was significantly associated with chronic HBV infection in one of the studies (24), but not in the other. One study reported decreasing HBsAg positivity rates in pregnant women according to study year, but not age (27). Regarding the health care workers, HBsAg prevalence was higher in older participants and considerably lower in central hospitals, indicating that there may be differences in awareness and/or safety policies at different levels of the health care system (35).

### Genotype distribution

We identified only 3 studies with HBV genotyping data in the Lao PDR (Table 3, Fig 2). The earliest study, published in 2008, reported a new genotype I (17). In another study, phylogenetic analyses of 42 HBV strains from HBsAg positive first time blood donors revealed mixed infections of sub-genotypes B1, B2, B4, C1, C5, I1 and I2 (18). The third study from 2014 reported strains belonging to genotype B, C and I (19). Besides B, C and I, no other genotypes were reported for the Lao PDR. In all three studies, genotypes B and C, or a mix of both, were found most often.

**Table 3.** Summary of the studies conducted on prevalence of HBV genotypes in the Lao PDR.

| Ref | Study location | Year of sample collection | study population | n | Genotypes |  |  |  |
| --- | --- | --- | --- | --- | --- | --- | --- | --- |
|  |  |  |  |  | B | C | I | Mixed (B, C, I) |
| (17) | VTN Capital, central provinces | NA | blood donors | 386 | 163 (42.2%) | 204 (55.4%) | 19 (4.9%) |  |
| (18) | VTN Capital | 2004-2005 | blood donors | 42 | 14 (33.3%) | 1 (2.4%) | 1 (2.4%) | 26 (61.9%) |
| (19) | VTN Capital, VTN Province, BLX | 2006 | blood donors | 43 | 19 (44.2%) | 21 (48.8%) | 3 (6.97%) |  |

VTN = Vientiane; BLX = Bolikhamxay Province; NA = not available; n = samples screened

## Discussion

This literature review provides an overview of studies investigating the genotype distribution and HBV serology in different subgroups and regions and highlights that our understanding of the epidemiology of Hepatitis B in the Lao PDR is incomplete. The aim of most of the studies was to estimate the rates of chronic infection in the Lao population; however, there are also some studies investigating the exposure to HBV, the level of protection conferred through vaccination against HBV or the genotype distribution.

Prevalence studies varied in study design and laboratory methods and included different population groups and therefore generalised statements about hepatitis epidemiology in the Lao PDR should be interpreted with caution. However, several important conclusions can be made, namely; high rates of chronic infection in adults, including women of child-bearing age; low infant vaccination coverage and compromised vaccine immunogenicity in particular regions; recent evidence of reduction in infection in adolescents born after vaccine introduction.

The earliest estimates of chronic HBV infection in the Lao PDR ranging from 8.7 to 9.6% were based on studies in blood donor samples derived from Vientiane Capital and central provinces in the years 2003 to 2006 (18–20). The classification of countries into high (≥8% HBsAg prevalence), intermediate (2-7%) and low (<2%) HBV endemicity is internationally recognized (38). Based on this, Lao PDR would be considered as “highly endemic”. However, while blood donors may represent an approximation for the general population, these studies were only conducted in central Lao PDR and do not provide any further information about other characteristics of the participants, such as for example ethnicity. In addition, the data may not mirror the current situation anymore as they are more than 14 years old.

Subsequent serological surveys provided further HBsAg estimates for women (ranging from 0 to 9.5%) and children and adolescents aged 8 months to 19 years (ranging from 0% to 11.8%) (12,20,23,25–29,31,32). One study estimated the rate of past infection for women at 40.3% (28). Drawbacks of these studies include non-randomized study design and the utilization of different testing methodology, which makes it difficult to compare the results. In addition, studies solely including women may underestimate the burden of hepatitis B. HBV screening of pregnant women and raising awareness in the general population are still very important public health interventions which need to be strengthened. Since MTCT is assumed to be the main route of transmission in the Lao PDR, it is unsurprising that maternal infection was identified as risk factor in two studies (23,24). Therefore, it is crucial to increase the coverage of the hepatitis B birth dose in order to reduce the transmission of hepatitis B in the Lao PDR. High infection rates represent a major public health challenge as treatment options in the country are still limited. Only 18% of patients in a retrospective cohort study conducted in 2018 describing the treatment of HBV infections in Vientiane Capital were reported to receive antiviral treatment (10).

Vaccination efforts will likely reduce the prevalence of chronic HBV in the Lao PDR in the long run, even though it is difficult to produce generalizable data on vaccine coverage and vaccine-induced protection due to the geographic and ethnic heterogeneity (which may play an important role) in the country (39). The aim of the World Health Organization - Western Pacific Region was to reduce HBsAg-prevalence to less than 1% in children aged 5 years and increase national coverage with both timely Hepatitis B birth dose (given within 24 hours of birth) and the third Hepatitis B dose to ≥95% by 2017 (40). At the moment, national coverage rates are at 55% and 85% for the birth dose and the third Hepatitis B dose, respectively (41). It is unlikely that the target of reducing the HBsAg prevalence under 1% in children below 5 years is met in the Lao PDR in the near future. To date, most of the data regarding the impact of vaccination are derived from studies that did not randomly select participants or only report the prevalence of HBsAg which hampers the possibility to extrapolate the data or to assess the levels of vaccine-induced protection. One cross-sectional randomized study reported decreasing exposure and infection rates after the introduction of HBV vaccination (32).

Despite the overall high prevalence of HBV infection in the Lao PDR, there are only two studies focusing on risk groups. In 2013, a high proportion of Lao health care staff was found to be susceptible to HBV and/or under-vaccinated (35). In addition, nearly half of the health care staff showed evidence of past exposure to the virus (48.9%). In the same year, a cross-sectional survey among students of health care professions was carried out in Vientiane Capital regarding their vaccination status, knowledge and awareness (42). Less than one third (21%) were fully vaccinated against HBV. When asked about the reason for not being vaccinated, the most common answer was that they did not know where to get vaccinated. At present, there is no national policy for the immunization or serological screening of health care workers, except for influenza vaccination. A clear occupational health vaccination policy is needed to protect health care workers and their patients and other at-risk populations. Furthermore, campaigns promoting vaccination and raising awareness of the risks for health care providers and their patients may prove beneficial.

In order to better protect potentially vulnerable populations, they must first be identified. Data are needed regarding HBV infection among sex workers and men who have sex with men. Furthermore, thalassemic and haemophilic patients and possible HBV co-infections with dengue, malaria or HIV are not yet investigated. The ethnic diversity in the Lao PDR poses another obstacle for the characterization of the HBV burden in the country. HBV infection rates among ethnic groups are not well described, apart from one study which took place in Saravan province(33) and could depend on local customs and risk practices such as tattooing, piercings, birth practices, sexual exposure etc.

The majority of the HBV strains characterized in the Lao PDR belonged to genotypes B and C. The distribution of genotypes found may have implications for the public health responses in the country, as genotype C was reported to show higher rates of progression from cirrhosis to hepatocellular carcinoma as compared to genotype B or other genotypes and showed a lower response rate to interferon (4,5).

In conclusion, Hepatitis B is still an important public health problem in the Lao PDR and more research is needed to better characterize its epidemiology. Interventions including HBV vaccination face many challenges but have showed first successes, at least in populations in central Lao PDR. In order to reduce the huge burden of HBV and its related morbidity and mortality in Lao PDR, such control measures need to be strengthened and sustained for the foreseeable future. Furthermore, the management of the existing HBV burden needs to be improved through increased testing and treatment capacities.

## Limitations

As a systematic review was performed, we did not include any existing grey literature on the topic. Study identification and data extraction were performed by one investigator only, possibly leading to selection bias. In addition, we cannot exclude the possibility that relevant articles were missed because they are not indexed in the databases used for the literature search. The heterogeneity of the studies concerning design, locations, methods and target population hampered the comparability of the findings.

## Data Availability

All data produced in the present work are contained in the manuscript

## Abbreviations

anti-HBs: Anti-hepatitis B surface antigen antibodies
anti-HBc: Anti-hepatitis B core antigen antibodies
DTPw-HepB-Hib: Diphtheria, tetanus, pertussis, hepatitis B and *Haemophilus influenzae* type b vaccine
HBsAg: Hepatitis B surface antigen
HBV: Hepatitis B virus
MTCT: Mother-to-child transmission

## Author contributions

**Lisa Hefele**: Conceptualization; Data curation; Formal analysis; Methodology; Validation; Visualization; Writing - original draft; Writing - review & editing

**Phonethipsavanh Nouanthong**: Methodology; Validation; Writing - review & editing

**Judith M. Hübschen**: Methodology; Validation; Writing - review & editing

**Claude P Muller**: Supervision; Methodology; Validation; Writing - review & editing

**Antony P Black**; Supervision; Methodology; Validation; Writing - original draft; Writing - review & editing

